# Motor evoked potentials as a side effect biomarker for deep brain stimulation programming

**DOI:** 10.1101/2025.01.24.25320924

**Authors:** Paola Testini, Austin Wang, Eric Cole, Svjetlana Miocinovic

## Abstract

**Objectives:** To determine if motor evoked potentials (mEP) – stimulation-induced muscle activation measured using electromyography – can serve as a biomarker of corticobulbar (CBT) and corticospinal (CST) tract activation for deep brain stimulation (DBS) programming.

**Methods:** In 12 patients with Parkinson’s disease and subthalamic or pallidal DBS, contact mapping determined clinical motor side effect thresholds. For equivalent stimulation parameters, EMG was recorded from cranial and arm muscles to determine the presence, peak amplitudes and latencies of mEP. Clinical and mEP thresholds were compared and accuracy metrics calculated to assess similarity between mEP and reported side effects.

**Results:** The mEP amplitudes increased with stimulation intensity. Latencies were shorter for cranial muscles, which were more likely to generate an mEP. Clinical and mEP thresholds were significantly correlated (R^2^ = 0.31; p=0.0006), although most mEP thresholds were lower than clinical side effect thresholds. The mEP accuracy in predicting side effects was 0.72, with a sensitivity of 0.68 and a specificity of 0.73.

**Conclusions:** EMG-recorded mEP correlated well with clinical side effects, and mEP often indicated subclinical CBT and CST activations.

**Significance:** This study characterizes motor potentials evoked by DBS and demonstrates their utility as an objective biomarker for motor side effect threshold detection during DBS programming.

**Highlights:**

- Deep brain stimulation can activate corticospinal/bulbar tract and evoke motor potentials in muscles measurable by surface EMG
- Motor evoked potential thresholds correlate significantly with clinical side effect thresholds but occur at lower stimulation intensities
- Motor evoked potentials may be a useful side effect biomarker for deep brain stimulation programming

## Introduction

Since its approval for tremor in 1997 and for Parkinson’s disease (PD) in 2002, deep brain stimulation (DBS) continues to be adopted for a growing number of indications (Lozano et al., 2019). The challenges and importance of programming for therapeutic success have been reported since early days (Krack et al., 2002; Volkmann et al., 2002). Patients undergo lengthy programming sessions with skilled programming clinicians to determine the best stimulation settings (contact configuration, stimulation amplitude, pulse width and frequency) to maximize therapeutic benefit and avoid side effects (Okun et al., 2008).

Establishing side effect thresholds (‘contact mapping’) is the first step in clinical DBS programming as it helps set allowable parameter ranges for further therapeutic exploration. Side effect thresholds are determined by gradually increasing stimulation amplitude while keeping other settings the same until a side effect is induced (this process is repeated for each contact configuration). Side effects occur when stimulation of the subthalamic nucleus (STN) or globus pallidus internus (GPi) causes inadvertent activation of the corticobulbar (CBT) or corticospinal (CST) tracts located in the internal capsule bordering the target nuclei, resulting in muscle spasms. Other types of side effects may also be induced by DBS such as sensory, visual, autonomic or affective, but motor side effects are typically the most common and therapy-limiting as they do not habituate. Clinical methods for determining side effect thresholds are relatively subjective as they rely on patient report and clinician observation. Patients can over-report perceived side effects as it can be difficult to differentiate muscle contractions due to CBT/CST stimulation from PD symptoms such as rigidity, dystonia, or pain (Mahlknecht et al., 2017). Similarly, patients may under-report side effects if CBT/CST activation is subtle, or if cognitive dysfunction or anxiety limits their active participation in the programming process.

In recent years, next-generation DBS devices and electrodes have allowed for more flexible and precise stimulation by increasing the number of stimulation contacts and expanding the parameter ranges (Falconer et al., 2018). However, this flexibility comes at a cost with each additional stimulation contact or parameter creating an exponential increase in the number of potential stimulation settings, leading to an even more time-consuming process difficult to perform during time-constrained clinical visits (Ten Brinke et al., 2018). As a result, there has been a strong interest in the DBS field to define objective biomarkers that would aid clinicians in selecting optimal stimulation settings more efficiently. Objective biomarkers could make the process less subjective or more accurate and reduce the entry barrier or training needed for programming (Cole and Miocinovic, 2025). Both imaging (Lange et al., 2021; Hines et al., 2024) and electrophysiologic (Binder et al., 2023; Busch et al., 2023; Lewis et al., 2024) biomarkers of therapeutic efficacy have been proposed. However, potential biomarkers for stimulation-induced side effects have received less attention (Cole and Miocinovic, 2025).

EMG responses from peripheral muscles have previously been studied in DBS patients (Ashby et al., 1999; Hanajima et al., 2004; Costa et al., 2007; Mahlknecht et al., 2017). It is well established that direct activation of CBT/CST can be elicited by DBS, and it may occur at stimulation amplitudes commonly used in clinical practice. DBS evokes ipsilateral and contralateral motor evoked potentials (mEP) measured with EMG in cranial and limb muscles, with likelihood increasing with stimulation amplitude(Costa et al., 2007). The physiologic explanation for this response is the axonal activation of the CBT/CST running in the posterior limb of the internal capsule near the DBS target nuclei, which then synapse to the neurons in the spinal cord and eventually lower motor neurons that innervate the muscles (Ashby et al., 1999; Costa et al., 2007; Mahlknecht et al., 2017; Miocinovic et al., 2018). The spread of stimulation effects to CBT/CST is related to the development of motor side effects such as muscle spasms in the face or limbs, or speech disturbances due to activation of laryngeal and tongue muscles (Volkmann et al., 2006; Tommasi et al., 2008; Mahlknecht et al., 2017). The mEP has been investigated as a tool for assisting in DBS lead placement intraoperatively, by providing a more accurate estimate of motor side effect thresholds (Nikolov et al., 2022; Trenado et al., 2024).

The goal of this study was to determine if mEP can be used as an objective biomarker of CBT/CST activation for future automated DBS programming applications. We hypothesized that mEP occur at the same stimulation settings that induce clinical motor side effects. We first systematically describe characteristics of mEP induced by STN and GPi DBS, including the latency and amplitude, which can inform a physiological understanding of mEP and guide their detection and therapeutic use. Next, we determine how mEP presence and mEP amplitude in cranial and limb muscles correlate with clinical motor side effects, noting high accuracy and concluding that mEP could provide a useful biomarker for side effect assessment in DBS programming.

## Materials and Methods

### 1. Patient selection

Patients with PD treated with STN or GPi DBS who had undergone stimulation for at least six months were recruited from the Emory Movement Disorder Center. The protocol was approved under the Institutional Review Board and the subjects signed an informed written consent.

### 2. Side effect testing and EMG recordings

The patients were evaluated in one research visit with dopaminergic medications withheld for 12 hours (medication off state). In patients with bilateral DBS, only one lead was stimulated. Patients underwent side effect threshold mapping of multiple DBS contact configurations (monopolar or bipolar stimulation, in a ring or pseudoring configuration) at a frequency of 130 Hz and a pulse width of 60 or 90 µs, similar to the standard clinical procedure. Threshold mapping was performed by increasing stimulation current amplitude, in 0.5 mA increments, until a motor or persistent sensory side effect was elicited (in which case motor side effect threshold was undefined). Side effects were determined by clinician’s (SM) visual observation or patient’s report of muscle contractions or speech changes, closely following clinical programming procedure, and categorized as resulting from activation of the CBT (facial contractions, vision or speech changes), CST (arm muscle contractions), or a combination of both tracts. Clinical motor thresholds were defined as the lowest stimulation amplitude required to elicit a motor side effect.

EMG signals were captured for mEP detection with Biosemi ActiveTwo system (Amsterdam, Netherlands) at a sampling rate of 16384 Hz (except one patient at 2048 Hz). EMG activity was recorded from nasalis, orbicularis oris, genioglossus, biceps, extensor carpi radialis (ECR), flexor carpi radialis (FCR), and first dorsal interosseus (FDI) muscles, although not all muscles were recorded in all patients (Table 1). Two flat surface EMG electrodes custom to the Biosemi ActiveTwo system were used for each muscle, spaced apart about 0.5 cm on the face and 1 cm on the arm. Cranial muscles were tested bilaterally since ipsilateral CBT innervates both sides of the head, while only contralateral arm muscles were tested since CST innervates primarily contralateral body.

**Table 1.** Patients’ clinical characteristics and experimental setup.

| Co<br>de | Age<br>range<br>(years)<br>/Sex | PD/DB<br>S<br>duratio<br>n<br>(years) | DBS<br>target/<br>Hemisph<br>ere<br>tested | Lead/IPG<br>model | Distal contact<br>coordinate (mm<br>from MCP) | #muscles<br>recorded<br>(contra.<br>face/ ipsi.<br>face/<br>contra.<br>limb) | #DBS<br>settings<br>tested<br>for<br>mEP/<br>side<br>effect | DBS<br>frequenc<br>y for<br>mEP/<br>side<br>effect | # DBS<br>contact<br>configuratio<br>ns with<br>amplitude<br>titration |
| --- | --- | --- | --- | --- | --- | --- | --- | --- | --- |
| Pt<br>1 | 60-69/F | 12/1.1 | STN/L | MDT 3389/<br>Activa SC | -9.3,-4.5,-5.9 | 2/2/2 | 25/20 | 30/130<br>Hz | 4<br>bipolar |
| Pt<br>2 | 60-69/M | 8/1.2 | STN/R | MDT 3389/<br>Activa RC | 12.0,-6.2,-3.9 | 1/0/1 | 40/22 | 30/130<br>Hz | 4<br>monopolar |
| Pt<br>3 | 60-69/M | 20/6.8 | STN/R | MDT 3389/<br>Activa SC | 11.0,-3.5,-5.0 | 2/2/2 | 30/24 | 30/130<br>Hz | 4<br>bipolar |
| Pt<br>4 | 50-59/M | 13/1.2 | STN/L | MDT 3389/<br>Activa RC | -11.6,-0.9,-1.6 | 2/2/1 | 24/21 | 30/130<br>Hz | 4<br>monopolar |
| Pt<br>5 | 60-69/M | 15/2.4 | STN/R | MDT 3389/<br>Activa RC | 8.6,-7.4,-8.4 | 2/2/2 | 26/22 | 30/130<br>Hz | 4<br>bipolar |
| Pt<br>6 | 60-69/M | 8/3.3 | STN/R | MDT 3389/<br>Activa RC | 12.1,-1.5,-3.6 | 2/2/2 | 41/24 | 30/130<br>Hz | 4<br>monopolar |
| Pt<br>7 | 50-59/M | 7/2.3 | STN/R | MDT 3389/<br>Activa RC | 10.9,-5.1,-2.5 | 2/2/2 | 62/46 | 30/130<br>Hz | 4 bipolar;<br>4<br>monopolar |
| Pt<br>8 | 50-59/M | 4/1.0 | STN/L | ABT 6172/<br>Infinity 5 | -14.0,-3.9,-3.6 | 2/0/2 | 110/61 | 10/130<br>Hz | 10<br>monopolar |
| Pt<br>9 | 40-49/M | 11/1.2 | STN/L | ABT 6172/<br>Infinity 7 | -10.7,-1.9,-5.1 | 2/0/3 | 79/50 | 10/130<br>Hz | 10<br>monopolar |
| Pt<br>10 | 40-49/M | 17/1.0 | STN/R | ABT 6172/<br>Infinity 7 | 9.8,-4.9,-5.9 | 1/0/2 | 50/29 | 20/130<br>Hz | 4<br>monopolar |
| Pt<br>11 | 70-79/M | 16/1.6 | GPI/R | MDT 3389/<br>Activa RC | 22.1,-0.3,-3.3 | 2/2/2 | 29/20 | 30/130<br>Hz | 4<br>monopolar |
| Pt 12 | 60-69/M | 11/1.9 | Gpi/L | MDT 3389/Activa SC | -21.2,2.0,-3.0 | 2/2/2 | 41/28 | 30/130 Hz | 4 monopolar |

IPG = implantable pulse generator; MCP = midcommissural point; contra = contralateral to stimulated hemisphere; ipsi = ipsilateral to stimulated hemisphere; mEP = motor evoked potential; MDT = Medtronic; ABT = Abbott.

EMG data was collected with the subject at rest in an armchair. Unilateral low-frequency (10-30 Hz) stimulation was applied at the same contact configurations and pulse widths as tested during high frequency stimulation, with amplitudes progressively increasing in 0.5 mA increments. Low frequency stimulation is required for mEP detection as the inter-pulse interval needs to be longer than the mEP latency to avoid overlap with the stimulation artifact. Because low frequency stimulation is not perceptible to patients, higher stimulation amplitudes than those used during high frequency stimulation were additionally applied to more completely characterize how mEP respond to stimulation. To minimize the effect of variable DBS contact impedances on the effects of stimulation, constant current stimulation was used. Constant current could be applied with Activa implanted pulse generators (Medtronic, Minneapolis, MN) at a stimulation frequency of 30Hz, while Infinity implantable pulse generators (Abbott, Plano, TX) allowed a stimulation frequency of 10 or 20 Hz. Approximately 5 seconds of data were recorded at each stimulation setting resulting in 50-150 applied DBS pulses.

### 3. Signal processing

EMG data was analyzed in MATLAB R2019b (MathWorks Inc.) with custom scripts. Signals were high pass filtered above 1 Hz using the 3rd order Butterworth filter. EMG data was epoched and averaged using the onset of the stimulation pulse as time zero. The presence or absence of mEP was determined by visual inspection by a single operator (PT) as a deflection from baseline that had consistent shape and latency across stimulation settings. The mEP shape and latency were generally consistent for the same patient and muscle across stimulation settings, allowing visual detection of even small deflections. If the determination was difficult, we used a consensus with two other investigators (SM and AW).

The mEP onset, peaks, troughs, and return to baseline were visually annotated using a MATLAB-based custom graphical user interface to precisely mark trough/peak points on the EMG trace at <0.1 ms accuracy. A maximum of five latency/amplitude points were selected. If the mEP was polyphasic and displayed more than three phases, lowest troughs and highest peaks were selected. The mEP amplitude was defined as the largest peak-to-trough difference in the mEP waveform and measured in microvolts (μV), and peak latency as the time interval from the beginning of the stimulation pulse artifact to the maximum peak of the mEP waveform, measured in milliseconds (ms). The mEP were categorized as resulting from activation of the CBT (if present in a cranial muscle), CST (if present in a limb muscle), or a combination of both tracts. The electrophysiological mEP threshold was defined as the lowest stimulation amplitude required to evoke an mEP in any muscle.

Since multiple muscles were recorded in each patient, we calculated a composite mEP value for each stimulation setting. Composite mEP was calculated by first normalizing each mEP amplitude from 0 to 1 (for each muscle and each patient separately) and then averaging normalized mEP across recorded muscles. Group recruitment curves were calculated by averaging composite mEP from different patients using the same active contact and same stimulation amplitude (at least two patients had to contribute to each data point in the group recruitment curves).

### 4. Clinical side effect and mEP comparison

To compare how well the presence or absence of mEP predicted clinical side effects, we considered clinical side effects to be the gold standard. Each stimulation setting where both EMG was recorded and clinical side effects were tested was classified as a true positive (presence of both mEP and side effect), false positive (presence of mEP and absence of side effect), true negative (absence of both mEP and side effect), or false negative (absence of mEP and presence of side effect). We then calculated accuracy, sensitivity, specificity, and F1 score using standard definitions.

Clinical and mEP thresholds were compared for each contact configuration that was tested at increasing stimulation amplitudes at both high and low stimulation frequency (stimulation amplitude titrations). For each contact configuration, we determined if clinical or mEP threshold was higher or lower or if only one measure had a defined threshold (e.g. if mEP was detected but there was no clinical side effect up to mEP threshold current, the configuration was classified as ‘mEP present, no side effect’). Indeterminate configurations were those where mEP threshold was above the highest stimulation amplitude tested for clinical side effects or vice versa, so it could not be determined which was higher or lower or if only one measure was present at the given threshold.

### 5. Statistics

Statistical analysis was performed in MATLAB. Descriptive results were presented as averages or medians. A Lilliefors test was used to test for normality. A rank sum Wilcoxon test was used to compare samples because mEP amplitudes and latencies were not normally distributed. Pearson linear regression coefficient was used to compare clinical and mEP thresholds. A chi-square test was used to determine which group of muscles was more likely to demonstrate mEP. A p-value of 0.05 or below was considered significant.

## Results

First, we summarize the patient population recruited for this study. Second, we show multiple examples of mEP recordings from different muscles and summarize their latency and amplitude characteristics, which can aid in understanding their physiological origin and inform how mEP can best be applied clinically. Next, we examine how accurately mEP can predict which stimulation settings produce clinical side-effects, noting high accuracy and that mEP appear at lower stimulation amplitudes. Last, we find that mEP presence is a superior predictor than mEP amplitude in differentiating how mEP relate to side effects.

### 1. Patient characteristics

Twelve patients with PD were included (1 female, 11 males) for a total of 12 hemispheres tested; 9 with STN and 2 with GPi DBS. Mean (±standard deviation) age at the time of testing was 59.7 (±8.8) years and patients had undergone DBS surgery 2.1 (±1.6) years earlier. A total of 557 stimulation settings were evaluated for mEP detection (range 24-110 per patient) and 367 stimulation settings evaluated for side effects (range 20-61 per patient). Across patients, a total of 64 contact configurations were tested at incremental stimulation amplitudes (‘amplitude titrations’). Clinical characteristics and experimental conditions are summarized in Table 1.

### 2. Motor evoked potential characteristics

Figure 1 displays examples of averaged EMG traces for different cranial and limb muscles at increasing stimulation amplitudes. In general, higher stimulation amplitude evoked a larger mEP once mEP threshold was reached. The average mEP peak latency was shorter for cranial muscles at 10.8±1.8 ms compared to limb muscles at 20.2±3.6 ms (p < 0.001; Fig. 2A). The mEP amplitude did not differ between the muscles (10.3±8.7 µV for cranial, 17.9±28.8 µV for limb; p = 0.16; Fig 2B). For cranial muscles, mEP oftentimes occurred during the falling phase of the electrical stimulation artifact so that the absolute peak-to-trough amplitude could not be reliably calculated, and instead relative amplitude was determined (Fig. 1).

**Figure 1.**
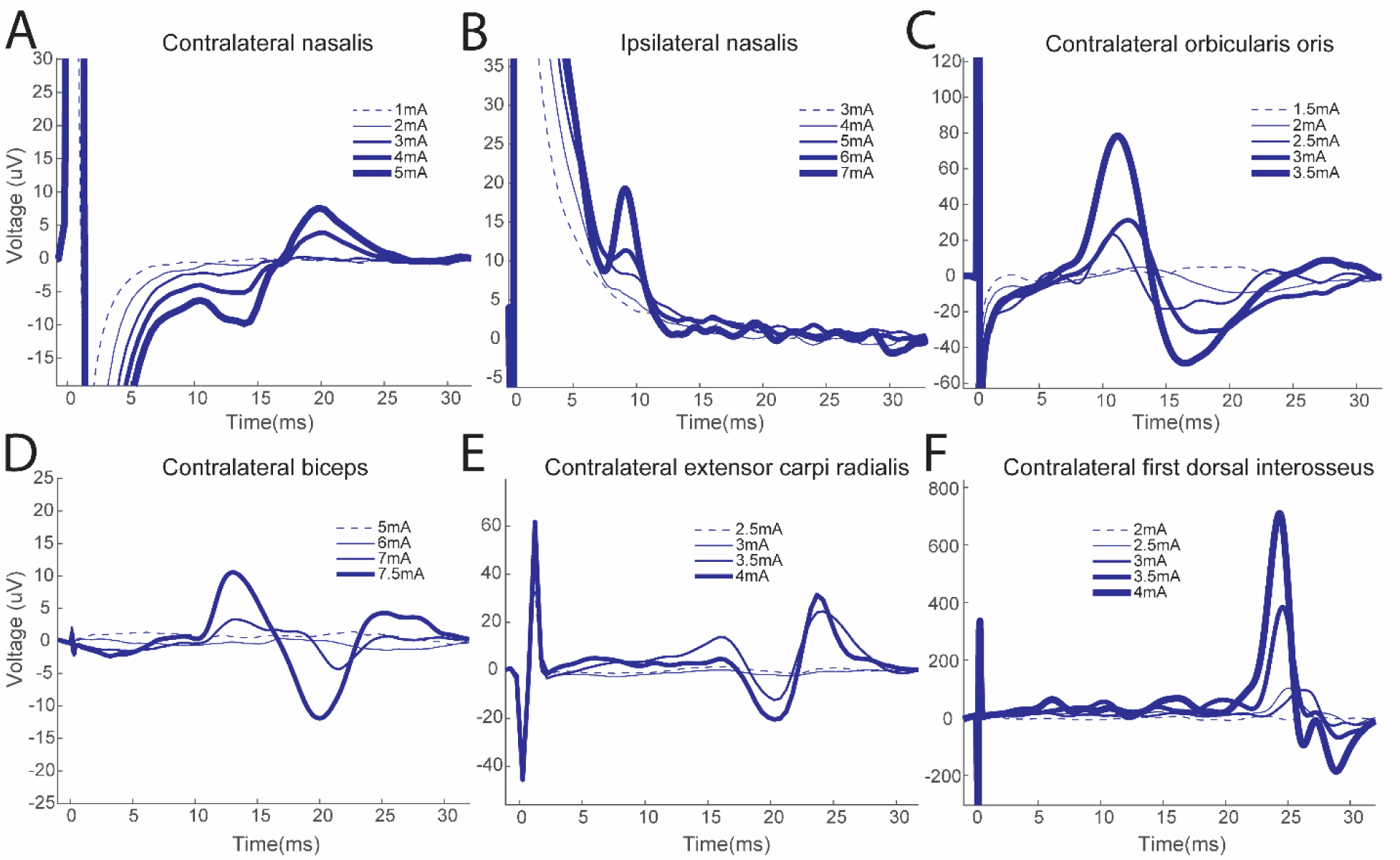
Examples of EMG traces (averaged by stimulation pulses at t=0) recorded in different muscles at increasing stimulation amplitudes in individual patients. For cranial muscles (top row), mEP can in some cases occur during electrical stimulation artifact. In limb muscles (bottom row), mEP occur at longer latencies.

**Figure 2.**
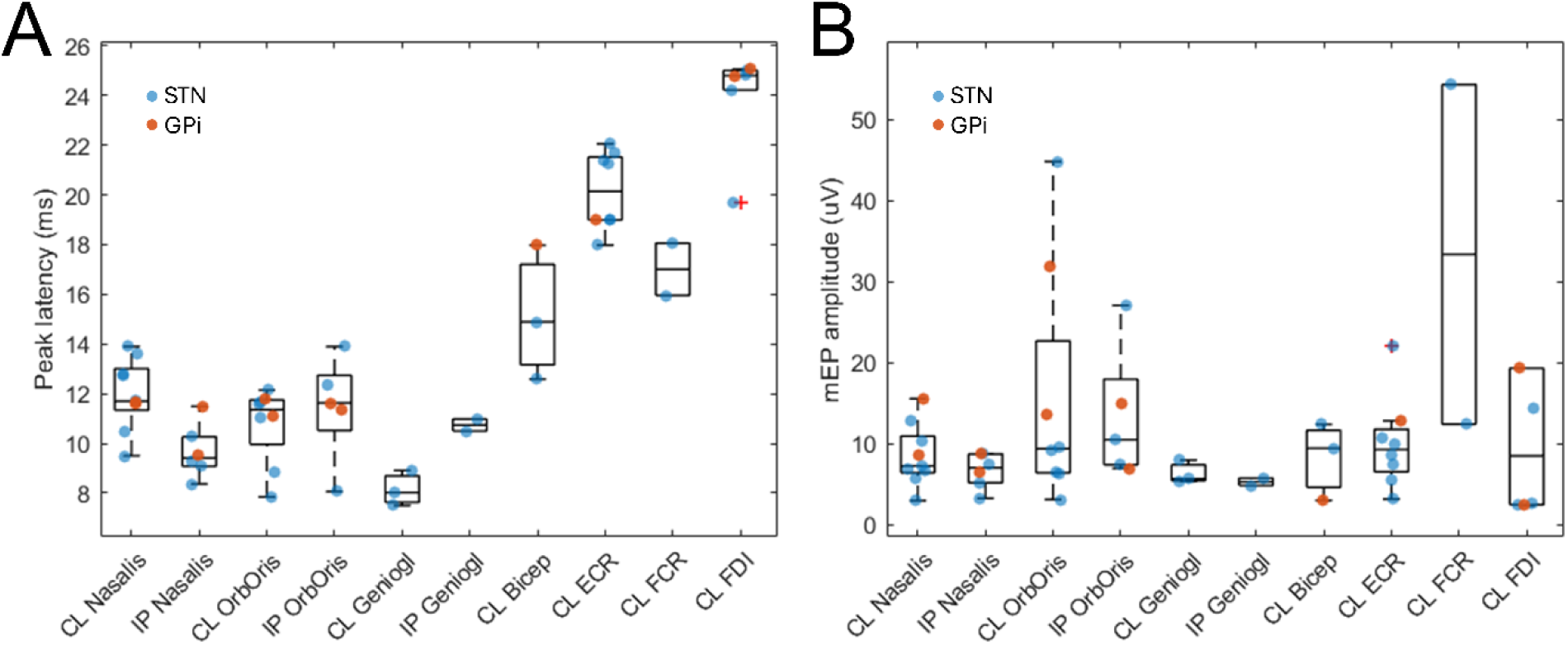
mEP peak latency (A) and peak-to-trough amplitude (B) by muscle. Each data point represents a patient median for each recorded muscle (blue = STN; red = GP). FDI amplitude has a blue outlier at 127 µV (not shown). CL = contralateral; IP = ipsilateral; OrbOris = orbicularis oris; Geniogl = genioglossus; ECR = extensor carpi radialis; FCR = flexor carpi radialis; FDI = first dorsal interosseus.

The mEP amplitude evoked from different contacts on the DBS electrode varied greatly between patients resulting in large error bars in group recruitment curves (Fig. 3). This was expected given that electrode position within local anatomy is known to differ across patient (Table 1). In general, mEP amplitudes increased with increasing stimulation amplitudes, but there were some exceptions observed in group data (Fig. 3) and individual patient recruitment curve (Fig. S1). Given the large variability between patients and small number of data points, statistical comparison was not performed on the recruitment curves. On visual observation, ventral contact (contact 0) stimulation led to larger mEP for both STN and GPi targets. When comparing monopolar and bipolar STN DBS, bipolar stimulation resulted in lower mEP amplitudes for the same stimulation current, and there was less difference in activation among contacts compared to monopolar stimulation.

**Figure 3.**
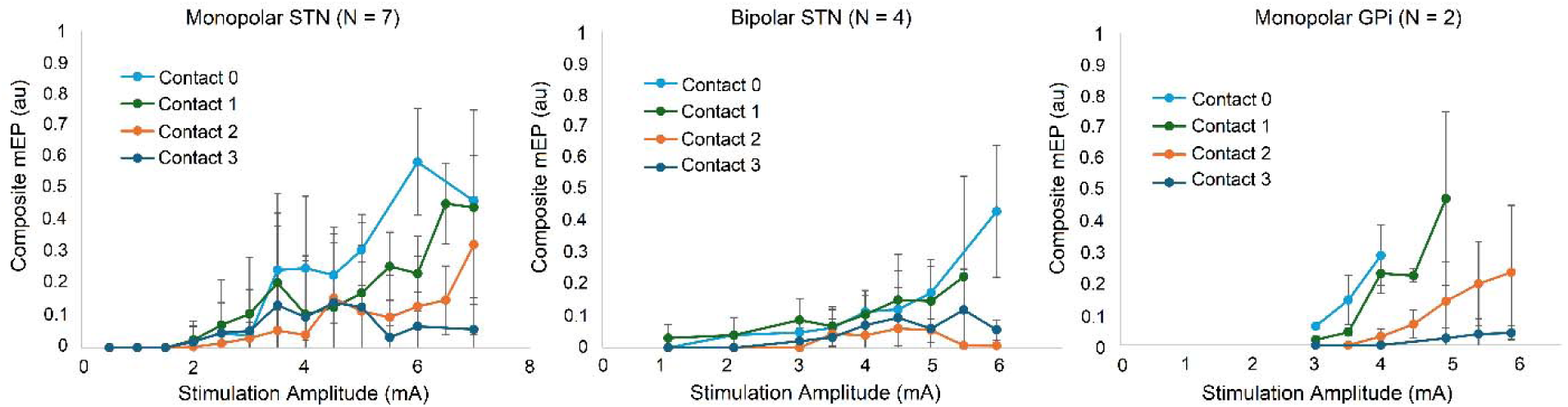
Recruitment curves for group data demonstrating change in composite mEP amplitude for each DBS target (STN or GP), stimulation configuration (monopolar or bipolar), active contact (0-3), and stimulation current (0.5-7mA). Pulse width for monopolar STN was 60 µs while bipolar STN and GPi were at 90 µs. Values from at least two patients were averaged to generate each data point. Error bars indicate standard deviation. Individual patient curves are available in the Supplementary section.

### 3. Comparison of motor evoked potentials and clinical motor side effects

The mEP accuracy in predicting side effects was 0.72, with a sensitivity of 0.68 and a specificity of 0.73. Due to the high number of true negatives, the F1 score was also calculated at 0.51 (Table 2). These metrics are affected by a high number of false positives (in most patients) and false negatives (majority from a single patient).

**Table 2.** Assessment of mEP as a marker of clinical motor side effect. Binary categorization of individual stimulation settings based on the presence or absence of mEP in any muscle (face or limb) and clinical motor side effect in any body part (corticobulbar or corticospinal). Side effect is considered the gold standard.

|  | TP (SE & mEP present) | TN (no SE & no mEP) | FP (no SE & mEP present) | FN (SE present & no mEP) | Accuracy | Sensitivity | Specificity | F1 score |
| --- | --- | --- | --- | --- | --- | --- | --- | --- |
| **All patients** | **54** | **211** | **77** | **25** | **0.72** | **0.68** | **0.73** | **0.51** |

| **combined** |  |  |  |  |  |  |  |  |
| --- | --- | --- | --- | --- | --- | --- | --- | --- |
| Pt 1 | 2 | 7 | 11 | 0 | 0.45 | 1 | 0.39 | 0.27 |
| Pt 2 | 6 | 12 | 4 | 0 | 0.82 | 1 | 0.75 | 0.75 |
| Pt 3 | 2 | 12 | 6 | 4 | 0.58 | 0.33 | 0.67 | 0.29 |
| Pt 4 | 5 | 10 | 6 | 0 | 0.71 | 1 | 0.63 | 0.63 |
| Pt 5 | 11 | 2 | 9 | 0 | 0.59 | 1 | 0.18 | 0.71 |
| Pt 6 | 5 | 16 | 3 | 0 | 0.88 | 1 | 0.84 | 0.77 |
| Pt 7 | 8 | 24 | 13 | 1 | 0.70 | 0.89 | 0.65 | 0.53 |
| Pt 8 | 0 | 43 | 0 | 18 | 0.70 | 0* | 1 | 0* |
| Pt 9 | 0 | 46 | 4 | 0 | 0.92 | NA* | 0.92 | 0* |
| Pt 10 | 4 | 20 | 3 | 2 | 0.83 | 0.67 | 0.87 | 0.62 |
| Pt 11 | 5 | 4 | 11 | 0 | 0.45 | 1 | 0.27 | 0.48 |
| Pt 12 | 6 | 15 | 7 | 0 | 0.75 | 1 | 0.68 | 0.63 |
| **Average** |  |  |  |  | **0.70** | **0.81** | **0.65** | **0.47** |

TP=true positive; TN = true negative; FP = false positive; FN = false negative; SE = side effect; mEP = motor evoked potential; NA = not applicable. \*Pt 8 had no recorded mEP but many reported side effects. Pt 9 had mEP but no reported side effects.

Because mEP occurred frequently without a side effect reported, we next investigated the relationship between mEP and side effect thresholds for all contact configurations where incremental amplitude titrations were performed (Fig. 4). In half of the configurations, mEP threshold was lower or equal to clinical threshold, while only in 3% mEP had a higher threshold (Fig. 4A). The majority of contact configurations in which an mEP was not recorded while a side effect was present (16%) came from a single patient. Overall, clinical and mEP thresholds were significantly correlated (R = 0.31; p=0.0006), and most mEP thresholds were lower than clinical side effect thresholds by 1.0±1.1 mA on average, and reflected in most data points lying above the unity line (Fig. 4B).

**Figure 4.**
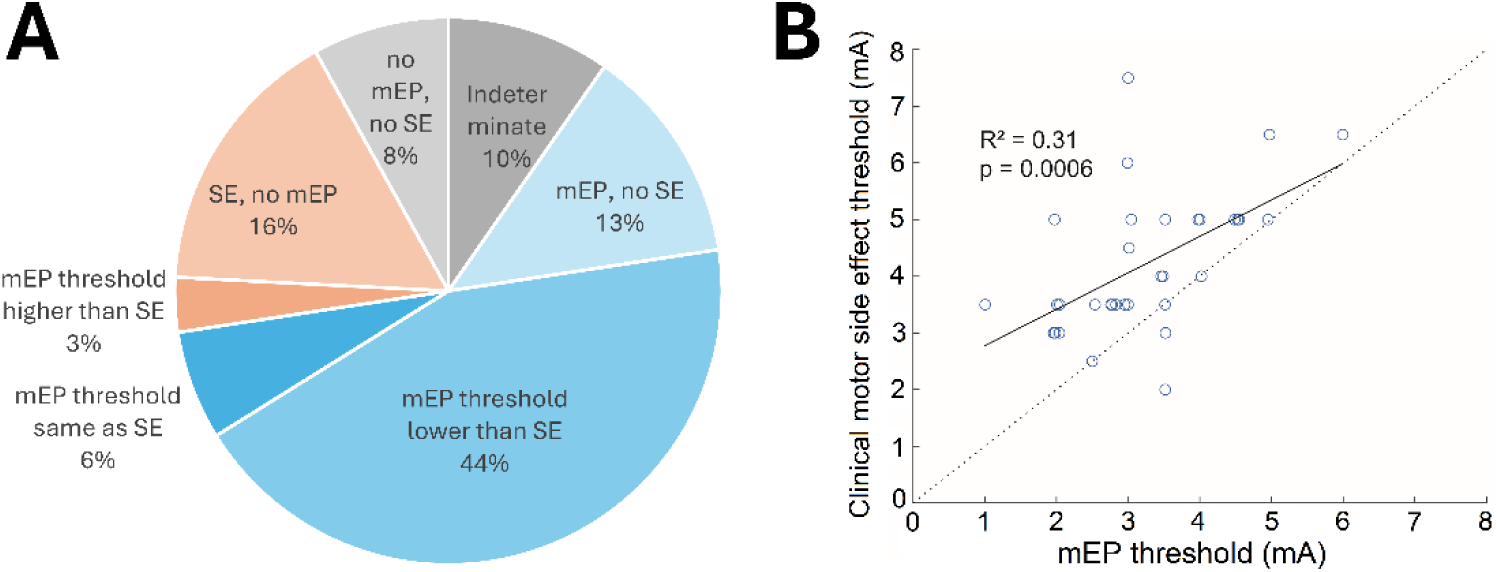
Threshold comparison for contact configurations with stimulation amplitude titrations (N=64). (A) Proportion of contact configurations based on their mEP and side effect (SE) occurrence and respective thresholds. (B) Comparison of mEP and side effect thresholds for configurations where both thresholds could be established (N=34). Side effect thresholds are mostly above the dashed unity line indicating that for a given contact configuration side effect threshold is higher than mEP threshold.

If a clinical side effect was reported, cranial muscles were more likely to show an mEP than limb muscles (chi-square; p<0.0001; Fig. 5). This was true even when side effects were separated by cranial or limb body distribution (Fig. S4). The genioglossus muscle was the most likely to demonstrate an mEP if a side effect was reported, but it was only recorded in 3 patients. Otherwise, the contralateral orbicularis oris and biceps muscles were the most likely to respond, for face and limb, respectively.

**Figure 5.**
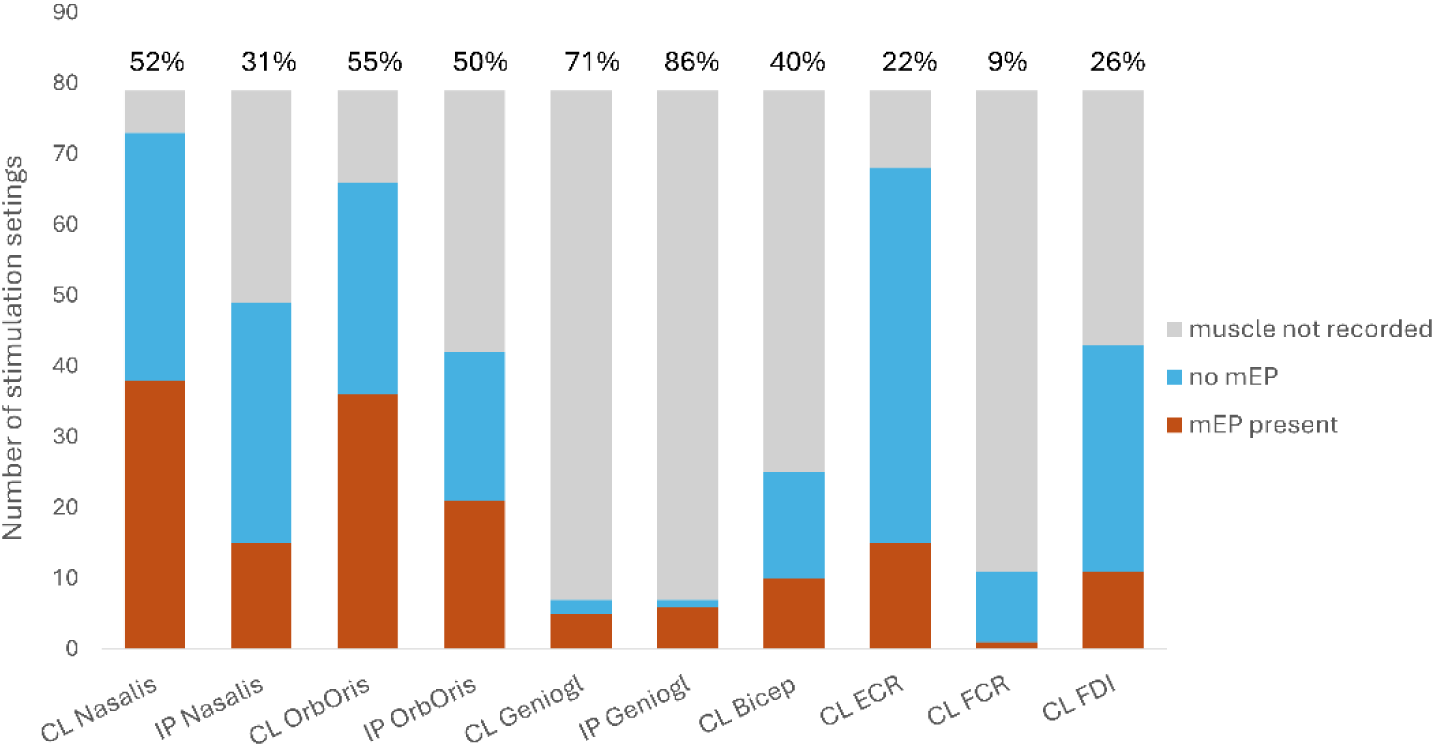
mEP response probability by muscle, only for stimulation settings that induced a side effect (N=79). Bars include settings for which muscles were not recorded to indicate the amount of data available, while numbers on the bars indicate proportion of stimulation settings for which a muscle was recorded and had an mEP. CL = contralateral; IP = ipsilateral; OrbOris = orbicularis oris; Geniogl = genioglossus; ECR = extensor carpi radialis; FCR = flexor carpi radialis; FDI = first dorsal interosseus.

We next tested whether the number of muscles with an mEP or mEP amplitude could predict presence of a clinical side effect for a given stimulation setting. For stimulation settings without a side effect, the median number of muscles with an mEP was 0 (interquartile range 1) and for those with a side effect it was 2 (interquartile range 3.75) which was significantly different (rank sum test, p<0.00001). The mEP amplitude was significantly higher in almost all muscles for stimulation settings which elicited a side effect compared to those that did not (Fig. S2A; the FCR muscle showed a trend toward significance with p=0.06). This was largely because for many stimulation settings that did not elicit side effects, the mEP was absent and therefore its amplitude was zero. When considering only mEP with non-zero amplitudes, this difference was not statistically significant except for the contralateral genioglossus (p=0.007). No difference was detected in latency of mEP when divided by presence or absence of side effect (Fig. S3). Therefore, more muscles showed an mEP when a side effect was present, but there was no mEP amplitude predictive of a side effect.

## Discussion

As the complexity of DBS programming increases and the indications for DBS expand, there is a growing need for objective biomarkers to guide programming. In this study, we correlated cranial and limb muscle mEP with side effects evoked by STN and GPi DBS in patient with PD, to evaluate mEP as a potential biomarker of CBT/CST activation for DBS programming. The mEP amplitudes varied by patient, and by stimulation contact, configuration and amplitude. The mEP peak latencies followed the expected anatomical distribution, with cranial muscles having shorter median latencies (range 8-12 ms) than arm muscles (range 15-25 ms). The mEP presence correlated significantly with presence of clinical motor side effects, although stimulation amplitude necessary to evoke mEP was usually lower than to evoke side effects, leading to false positives (presence of mEP without side effects) and affecting accuracy and specificity metrics.

### Anatomical insights from mEP measurements

Our findings on characteristics of mEP agree with prior studies (Ashby et al., 1999; Costa et al., 2007; Tommasi et al., 2008; Mahlknecht et al., 2017). The latency of mEP varied by muscle; proximal muscles had shorter latencies while distal muscles had longer latencies, consistent with the distance travelled by the evoked action potentials from the site of activation in the brain to the neuromuscular junction of individual muscles. The amplitude of the mEP increased with increasing stimulation amplitudes suggesting greater CBT/CST recruitment. This result is consistent with CBT/CST potentials evoked by DBS that were measured at the cortical surface (Miocinovic et al., 2018). Cranial muscles were more likely to generate mEP than limb muscles for stimulation settings which elicited side effects, regardless of the location of the side effects, consistent with the observation that cranial motor side effects tend to occur at lower stimulation amplitudes than limb motor side effects(Tommasi et al., 2008). This is likely related to the somatotopic organization of CBT and CST. The two tracts travel jointly in the internal capsule as they pass the STN and GPi on the way to the brain stem and the spinal cord. CST axons synapse onto motor neurons in the spinal cord that innervate contralateral trunk and limb muscles, while CBT axons synapse in cranial nerve nuclei in the brain stem which innervate bilateral facial, tongue and laryngeal muscles(Davidoff, 1990; Morel et al., 1997). The studies on somatotopic organization indicate that CBT descends through the anterior part of the posterior limb of the internal capsule, with face-arm-leg fibers further organized in antero-posterior and medio-lateral axes meaning that the face is the most anterior and medial(Bertrand et al., 1965; Duerden et al., 2011). The STN is located medially to the internal capsule while GPi is lateral to it. The proposed somatotopic organization explains why STN DBS activates bulbar muscles before limb muscles. However, in our two patients with GPi stimulation, cranial muscles also had lower mEP thresholds compared to limb which suggests that there may be considerable overlap in tract trajectories.

With a few exceptions, mEP amplitudes were higher with more ventral stimulation contacts, suggesting a closer proximity to the internal capsule for the ventral contact. In clinical programming for STN implantations, the most dorsal contact is often thought to be anatomically closest to the internal capsule based on STN location and electrode angle (Marks, 2015). Our finding showing largest mEP ventrally is consistent with prior mEP studies (Ashby et al., 1999; Mahlknecht et al., 2017) and highlights the fact that the most ventral STN contact significantly contributes to CBT/CST activation which should be considered during clinical programming. For GPi implants, the proximity of the most ventral contact to the internal capsule is expected since internal capsule courses posterior (and medial) to the nucleus (Marks, 2015) which is consistent with our mEP findings. Larger CBT/CST evoked potentials measured in the cortex were also found with stimulation of the most ventral contacts in STN and GPi in a previous study (Miocinovic et al., 2018). Detailed anatomical reconstructions of contact locations and estimations of DBS-induced electrical field were outside the scope of this study and will be addressed in future work.

### Relationship between mEP and clinical motor side effects

The presence of mEP correlated well with presence of clinical motor side effects in all but one patient, suggesting that mEP could be a useful biomarker of side effects for DBS programming. The outlier patient reported motor side effects at nearly all contact configurations, despite lack of mEP even at much higher stimulation amplitudes than side effect thresholds. There are several possible reasons: 1) appropriate muscles were not recorded (e.g. in this patient ipsilateral cranial muscles were omitted); 2) patient may have mistaken sensory paresthesia for muscles contractions, but the patient’s lateral lead location makes this less likely; 3) patient was the only subject who fell asleep during mEP recordings which may have affected CBT/CST activations or EMG electrode skin adherence. Another patient reported no motor side effects, but this was appropriate as mEP occurred at high stimulation amplitudes, which led to paresthesia and nausea during high stimulation frequency. These examples illustrate the fact that patient-reported side effects may not always be reliable. Some patients may not know what to expect and give reports during programming that are misleading or inaccurate. Thus, it’s important to acknowledge that the traditional method of assessing side effects during programming is not a perfect ground-truth, which may have affected the results of this study and made mEP look like a less accurate biomarker than it could be. Such an effect would be very challenging, if not impossible, to measure empirically, and further emphasizes the need for objective biomarkers to guide programming.

We had to use low frequency stimulation as a surrogate marker of high frequency activation, as the latter would limit EMG signal interpretation due to DBS artifact. The mismatch in mEP and clinical thresholds could occur due to differences in spatial spread of stimulation for low versus high frequency stimulation, but we think this is unlikely. First, during low frequency stimulation, patients are unable to perceive muscle contractions even at large stimulation amplitudes that elicit clear mEP in multiple muscles. This suggests that perception of a muscle spasm requires that the muscle tonically contracts which happens during temporal summation of high frequency action potentials. Second, computational DBS activation models consistently demonstrate that differences in stimulation frequency do not affect spatial spread of stimulation, but instead alter temporal dynamics of neural firing (McIntyre et al., 2004). Nonetheless, to investigate the role of frequency effect, future studies should compare mEP at different stimulation frequencies.

In our study, mEP often indicated subclinical activation of CBT/CST because they occurred at lower stimulation amplitudes where patients did not report and clinician did not observe motor side effects. This suggests that patients and clinicians might frequently be unaware when DBS effects first start spreading into the CBT/CST. While lack of perceived side effects may suggest that internal capsule activation is not clinically relevant, CBT and CST stimulation may lead to side effects after chronic stimulation (Tommasi et al., 2008). The mEP thresholds can also be lowered by voluntary activation of the same or different muscles which can potentially lead to overestimation of motor thresholds during contact mapping and occurrence of motor side effects outside of the clinic in patients’ daily life (Mahlknecht et al., 2017). Subclinical CBT/CST activations also may help explain why many patients experience speech or gait difficulties after DBS (Phokaewvarangkul et al., 2019; Tripathi et al., 2024). This is another important question that should be further investigated in future studies.

The main limitations of our study are: 1) the small number of patients, particularly for the GPi target; 2) the variability in number of muscles assessed; 3) the lack of recordings from the lower limb (in test recordings, leg mEP were not observed); 4) the variability in stimulation parameters used across patients which was partially due to time constraints that limited our ability to perform complete amplitude sweeps at multiple contact configurations; 5) mEP detection was performed visually, which may have introduced subjectivity; however, this was performed by a single rater limiting variability and the rater was blinded to clinical side effect thresholds. Development of automated mEP detection signal processing methods would be a valuable addition to future studies; 6) cranial mEP exhibited shorter latencies and therefore sometimes superimposed on the stimulation artifact, limiting some absolute mEP amplitude calculations; however, this did not affect the relative comparison of mEP amplitudes elicited by different stimulation amplitudes because within the same patient and contact configuration, the same muscle always had similar stimulation artifact; 7) in some instances a higher stimulation amplitude did not generate an mEP or generated mEP of smaller amplitude compared to lower current, typically with the higher stimulation intensities. The physiological reason for this effect is unclear, though it could result from muscle fatiguability (which our study was not designed to evaluate). Reliability of measurements could have been improved by repeating the same stimulation setting more than once but this was not done due to experimental time limitations; 8) finally, only patients with PD were included, possibly limiting the generalizability of our results.

## Conclusions

Using surface EMG to assess motor evoked potentials, CBT and CST activation were detected at lower thresholds than clinical determination of motor side effects, with cranial muscles more frequently predicting side effects than limb muscles. These results suggest that EMG, particularly when recorded from bulbar muscles, is a potential biomarker for objective STN and GPi DBS programming. Its activation at lower thresholds may protect from overestimation of motor thresholds, leading to fewer side effects after chronic stimulation or stimulation during motor activity. EMG-recorded mEP is a potentially useful biomarker for DBS programming. The potential benefits include faster and more accurate assessment of motor side effect thresholds and enabling future applications for automated programming alongside optimization algorithms and therapeutic biomarkers. The potential importance of subclinical CBT/CST activation and optimal stimulation frequency for mEP detection should be evaluated in future studies.

## Data Availability

All data produced in the present study are available upon reasonable request to the authors after final publication.

## Supplementary Figures

**Figure S1.**
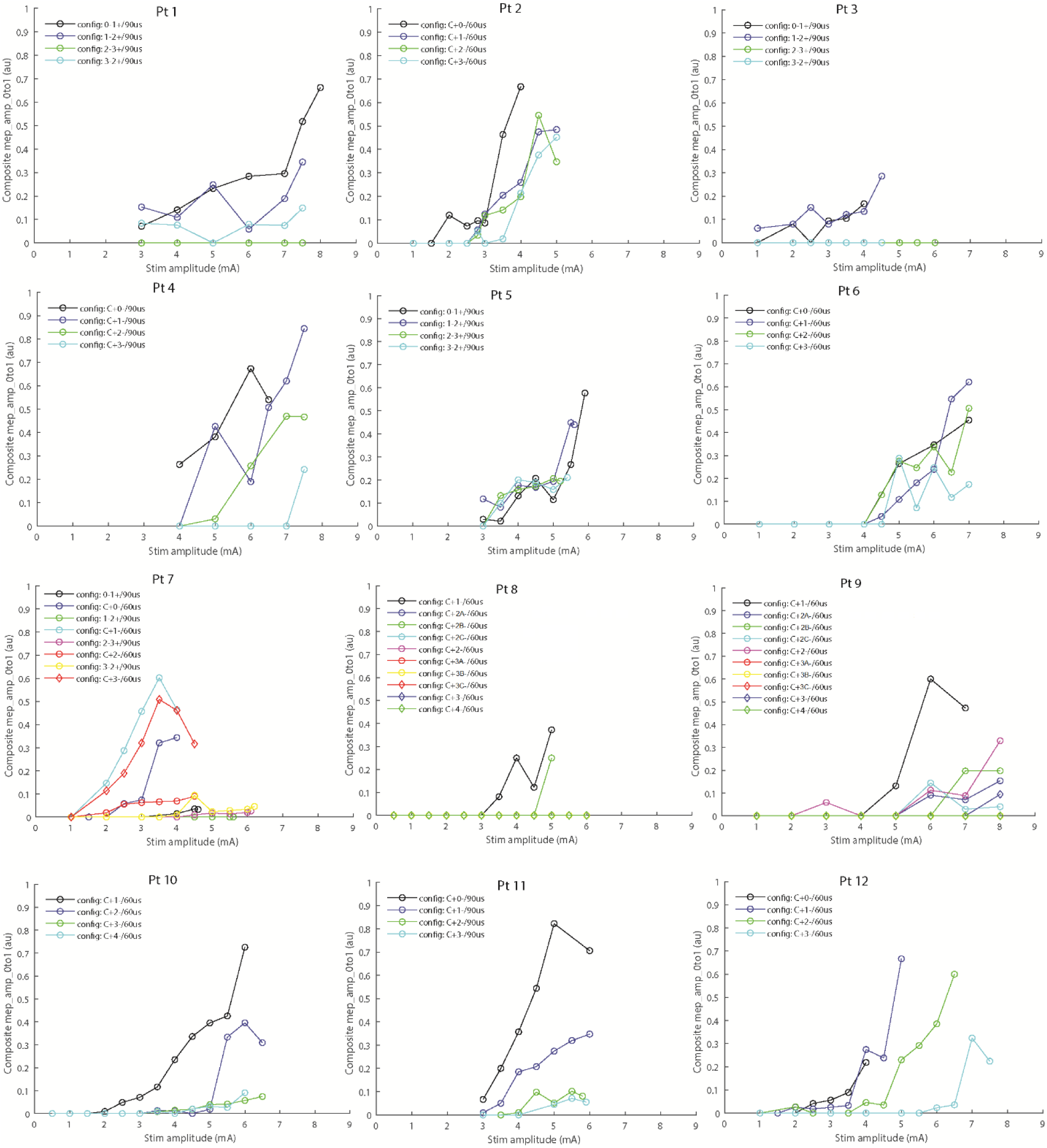
Recruitment curves for individual patients. mEP composite amplitude is created by normalizing each mEP amplitude from 0 to 1 (for each patient and each muscle), then averaging across all muscles so that each stimulation setting is associated with one composite mEP amplitude.

**Figure S2.**
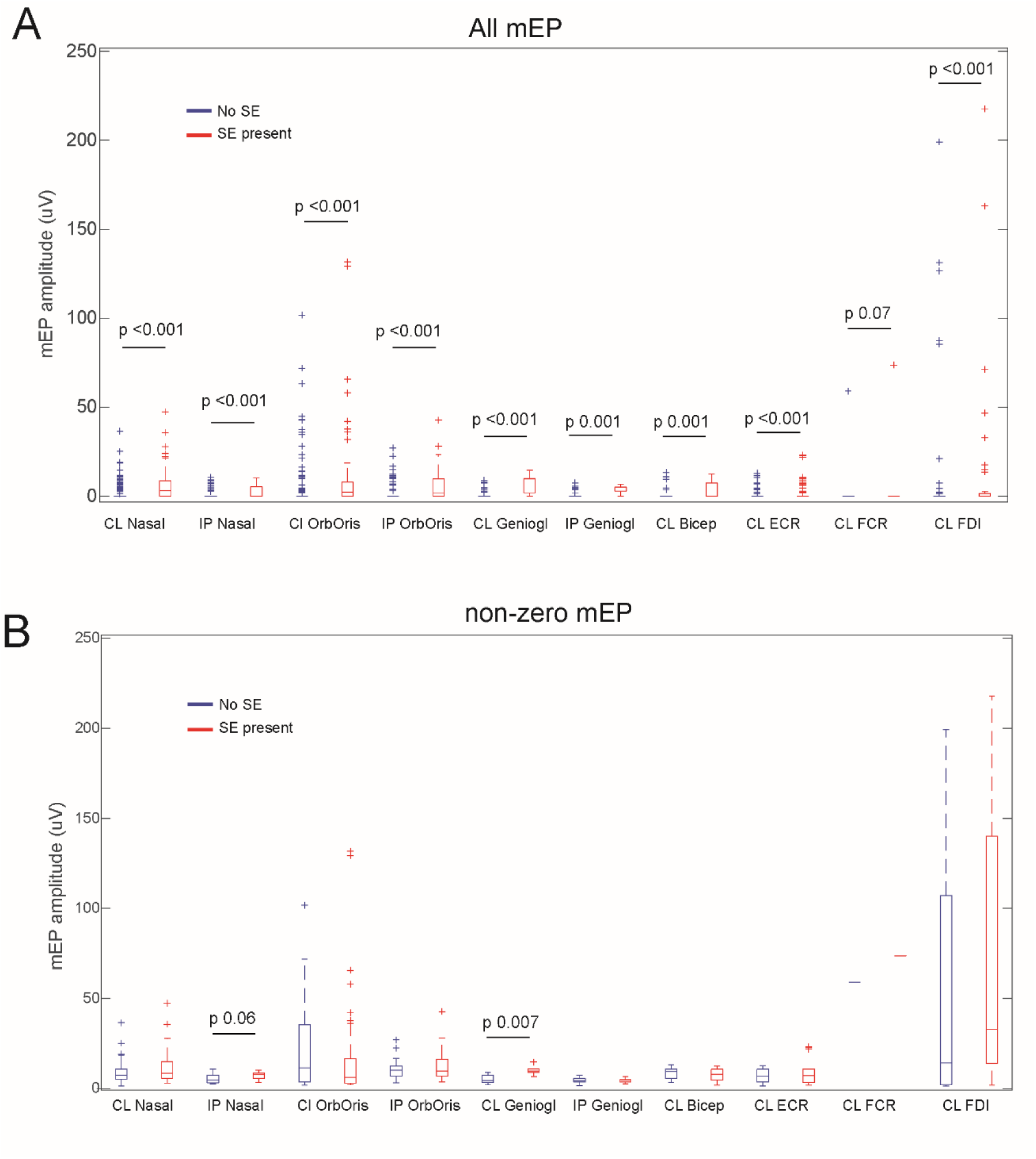
mEP amplitudes for all patients and all muscles, divided based on association with a clinical side effect (SE). On each box, the central mark indicates the median, and the bottom and top edges of the box indicate the 25th and 75th percentiles; the whiskers extend to the most extreme data points not considered outliers, and the outliers are plotted using the ‘+’ symbol. The last boxplot has an outlier at 476uV which is not shown. The last boxplot has an outlier at 476uV which is not shown. (A) mEP amplitudes for all settings are plotted (including mEP=0). (B) only non-zero mEP are plotted. CL = contralateral; IP = ipsilateral; OrbOris = orbicularis oris; Geniogl = genioglossus; ECR = extensor carpi radialis; FCR = flexor carpi radialis; FDI = first dorsal interosseus.

**Figure S3.**
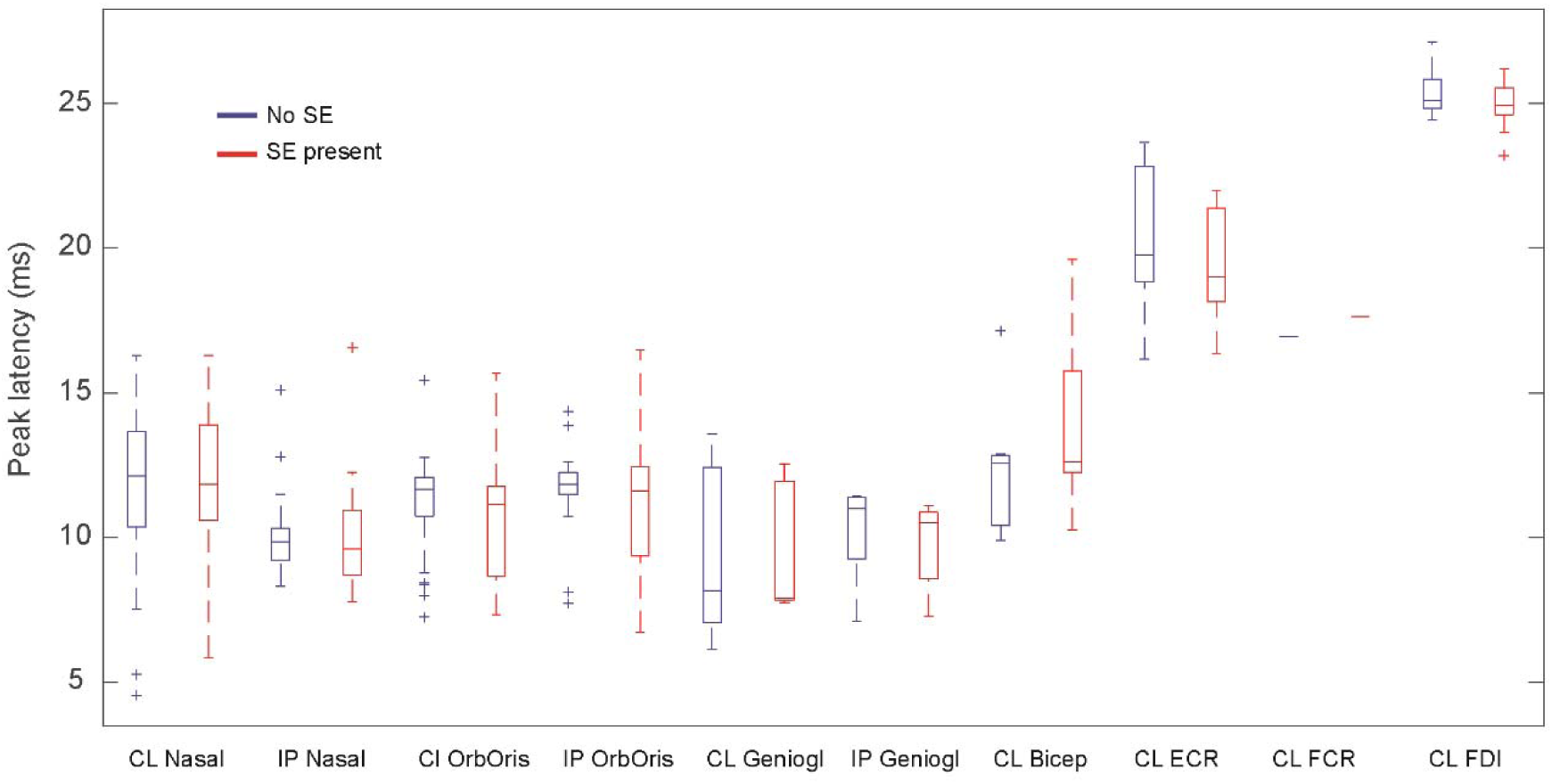
mEP latencies for all patients, divided based on association with a clinical side effect (SE). On each box, the central mark indicates the median, and the bottom and top edges of the box indicate the 25th and 75th percentiles; the whiskers extend to the most extreme data points not considered outliers, and the outliers are plotted using the ‘+’ symbol. P-values from Wilcoxon rank sum test were non-significant. CL = contralateral; IP = ipsilateral; OrbOris = orbicularis oris; Geniogl = genioglossus; ECR = extensor carpi radialis; FCR = flexor carpi radialis; FDI = first dorsal interosseus.

**Figure S4.**
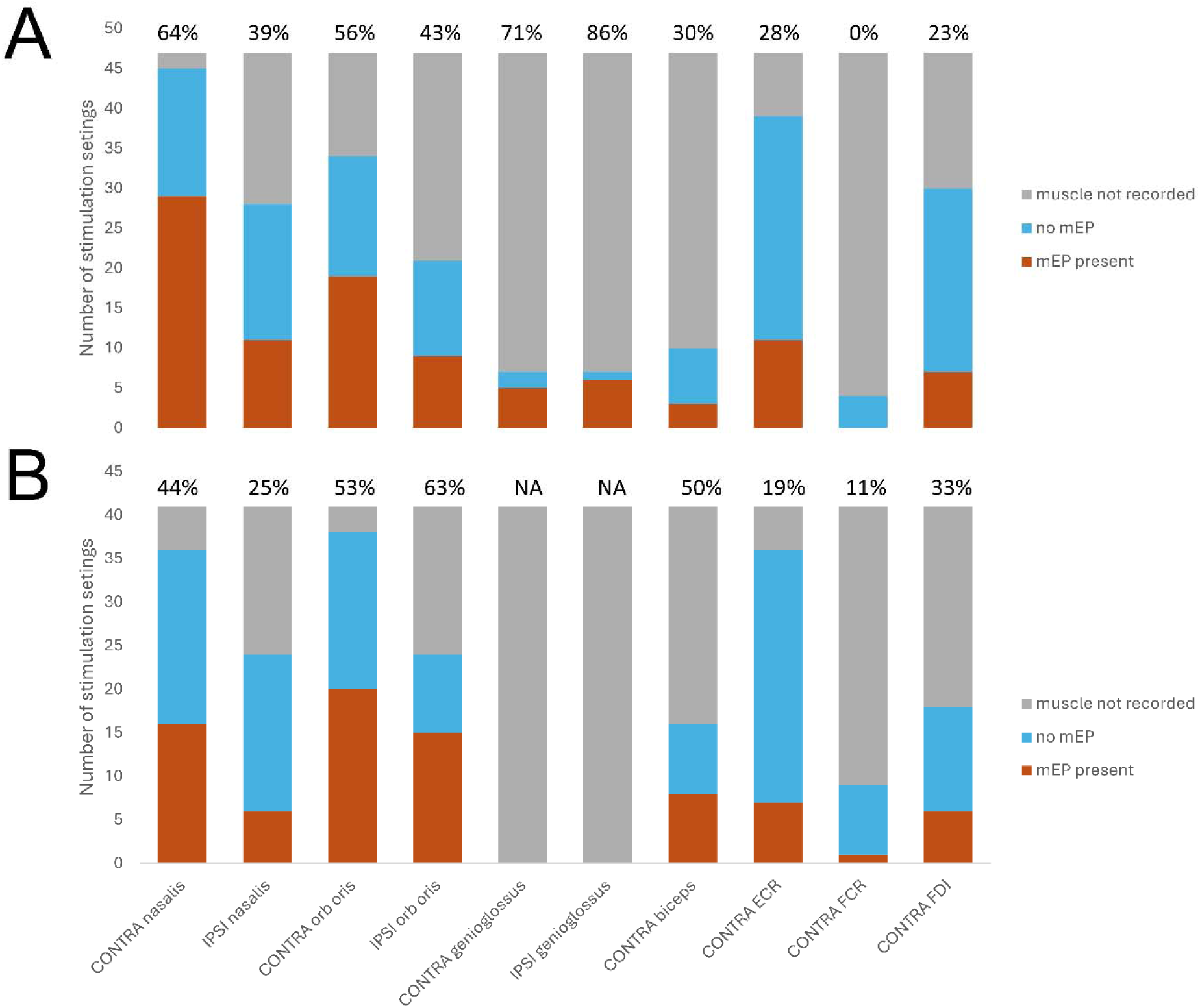
mEP response probability by muscle, only for stimulation settings that induced (A) a corticobulbar side effect (N=47); or (B) a corticospinal side effect (N=41). Bars include settings for which muscles were not recorded to indicate the amount of data available, while numbers on the bars indicate proportion of stimulation settings for which a muscle was recorded and had an mEP. CL = contralateral; IP = ipsilateral; OrbOris = orbicularis oris; Geniogl = genioglossus; ECR = extensor carpi radialis; FCR = flexor carpi radialis; FDI = first dorsal interosseus.

